# Effect of hydroxychloroquine on SARS-CoV-2 viral load in patients with COVID-19

**DOI:** 10.1101/2020.06.16.20133066

**Authors:** Klinger Soares Faíco-Filho, Danielle Dias Conte, Luciano Kleber de Souza Luna, Joseane Mayara Almeida Carvalho, Ana Helena Sitta Perosa, Nancy Bellei

**Affiliations:** Universidade Federal de São Paulo; Universidade Federal de São Paulo, Hospital São Paulo

**Keywords:** viral load, SARS-CoV-2, hydroxychloroquine, COVID-19

## Abstract

**Background:** Some studies have shown that hydroxychloroquine (HCQ) is an effective drug in reducing the *in vitro* replication of SARS-CoV-2. However, the *in vivo* effect of HCQ still unclear. This study aims to evaluate viral load clearance in patients with COVID-19 who underwent HCQ treatment in comparison with a control group that did not receive the drug.

**Methods:** This prospective study comprised consecutive viral load measurements in patients with COVID-19 hospitalized with a moderate illness. Patients received 400 mg of HCQ every 12 hours for 10 days according to the medical decision. Nasal swab samples were collected at the 1^st^, 7^th,^ and 14^th^ days of the admission.

**Results:** 155 samples were collected from 66 patients with COVID-19 (60% female), with a median age of 58 years. The viral load between studied groups, assumed as a semiquantitative measure of cycle threshold (Ct) values, presented no significant difference within the three consecutive measures (ΔCt) (*p>0*.*05*). We also analyzed the ΔCt viral load at different intervals of sample collection (Δt <7; 7-12 and >12 days) without significant differences at any ΔCt (*p>0*.*05*).

**Conclusion:** In this study, we did not observe any change in viral load *in vivo* with the use of HCQ.

**Summary:** We evaluate viral load clearance in patients with COVID-19 who took hydroxychloroquine (HCQ) for treatment and those who not. Prospective viral load measurements have shown any change in viral load in vivo with the use of HCQ.

## INTRODUCTION

On March 11, 2020, the World Health Organization (WHO) declared the novel severe acute respiratory syndrome coronavirus (SARS-CoV-2), responsible for the coronavirus disease of 2019 (COVID-19), a pandemic, when the virus reached five continents [1]. Since then, several medications have been tested in the treatment of this disease such as hydroxychloroquine (HCQ) [2], tocilizumab [3, 4], remdesivir [5], and heparin [6].

Some studies have already demonstrated the broad-spectrum antiviral potential of HCQ, a drug widely used as an antimalarial or in the treatment of autoimmune disease [7, 8]. Some studies have demonstrated the *in vitro* effectiveness of HCQ and chloroquine in controlling the replication of SARS-CoV-2 [9, 10].

On May 22, a multinational registry analysis was published with 96032 patients with COVID-19 that received chloroquine alone or combined with a macrolide, as well as HCQ alone or combined with a macrolide. The study found that hospitalized COVID-19 patients treated with HCQ had a higher risk of death than those in the control group [11]. As a result, WHO advised against continuing studies with HCQ as an alternative drug for the treatment of COVID-19. However, several concerns were raised regarding the veracity and analysis of the data conducted by the authors. Due to concerns involving reevaluation of original data by a new peer review, the authors could no longer vouch for the veracity of the data and requested the retraction of the paper [12]. Therefore, more studies need to be carried out to understand the real benefit of HCQ in the treatment of the disease. The present study aimed to evaluate viral load clearance in patients with COVID-19 who underwent treatment with HCQ in comparison with a control group that did not receive the drug.

## MATERIAL AND METHODS

### Patients and drug administration

This prospective study comprised consecutive viral load evaluations in patients with COVID-19. A total of 155 samples from 66 hospitalized patients at university hospital Sao Paulo, Brazil, aging > 18 years of age, were included in the study. They were diagnosed with severe acute respiratory syndrome (SARS) due to COVID-19. We excluded patients under 18 years old, ICU patients, and those presenting with severe conditions including malignancy, heart, liver, or renal diseases and severe decompensation (estimated glomerular filtration rate ≤ 30 mL/min/1.73 m^2^), inadequacy for oral administration, inability to cooperate due to cognitive impairment or poor mental status, pregnancy or lactation and HCQ allergy. In the original protocol, patients with severe COVID-19 were excluded.

The HCQ was prescribed according to medical decision and acceptance of the patient. A dose of 400 mg HCQ was administered every 12 hours for 10 days. The patients were included after approval of the study by the Hospital Research Ethics Committee (CEP n. 4.013.602).

### Samples and RNA preparation

Nasal swab samples were collected from patients during the early (Ct1), intermediary (Ct2), and final clinical stage of COVID-19 (Ct3), corresponding to the 1st, 7th, and 14th days of the admission, respectively, to evaluate the viral clearance.

The RNA of samples was purified using the Quick-RNA Viral Kit (Zymo Research, USA) according to manufacturer instructions. Purified RNA was stored at -80°C.

### SARS-CoV-2 detection

Viral detection was performed with AgPath-ID One-Step RT-PCR Reagents, in a total of 20 µL reaction volume, containing 5,0 µL of purified RNA, primers and probes (400 nM and 200 nM, respectively) aiming at the CDC USA protocol N1 and N2 targets of the SARS-CoV-2 nucleoprotein gene, and human ribonuclease P gene (RNAse P) as endogenous control [13]. Samples with Ct values < 40 were considered positive.

### Viral load analysis method

We used the cycle threshold (Ct) values as a semiquantitative measure of viral load. The amount of viral RNA target present in positive samples is inversely proportional to the corresponding Ct value, meaning that the greater amount of viral RNA, the lower the Ct value obtained. To assess the Ct value variation along time, consecutive RT-PCR tests were conducted in the patients with COVID-19, during hospitalization or after discharge, until undetected Ct values. The variation between consecutive Ct values for each patient analyzed was evaluated as ΔCt.

### Statistical analysis

Statistical analysis was performed using the chi-square test (χ^2^) and two-way ANOVA to compare categorical values, with a significance level of p <0.05.

## RESULTS

Among the 66 patients with laboratory-confirmed COVID-19, admitted at São Paulo Hospital, 60% (40/66) were male. The median age was 58 (range 18-85) years. The patients presented mild to severe symptoms, radiographic evidence of pneumonia, feverish or not, and received supplemental oxygen therapy until 5 L/min and, but none were classified as ‘severe’ or ‘critical’, and none were further admitted to ICU. The majority of patients reported comorbidities such as hypertension, diabetes mellitus type 2, and dyslipidemia. The 155 nasal swabs from the 66 patients were collected from first-day symptoms up to the 36th day. We detected the highest viral loads soon after symptom onset, which then gradually decreased towards the detection limit at about the second week. Viral loads were very heterogeneous as shown in Figure 1.

**Figure 1:**
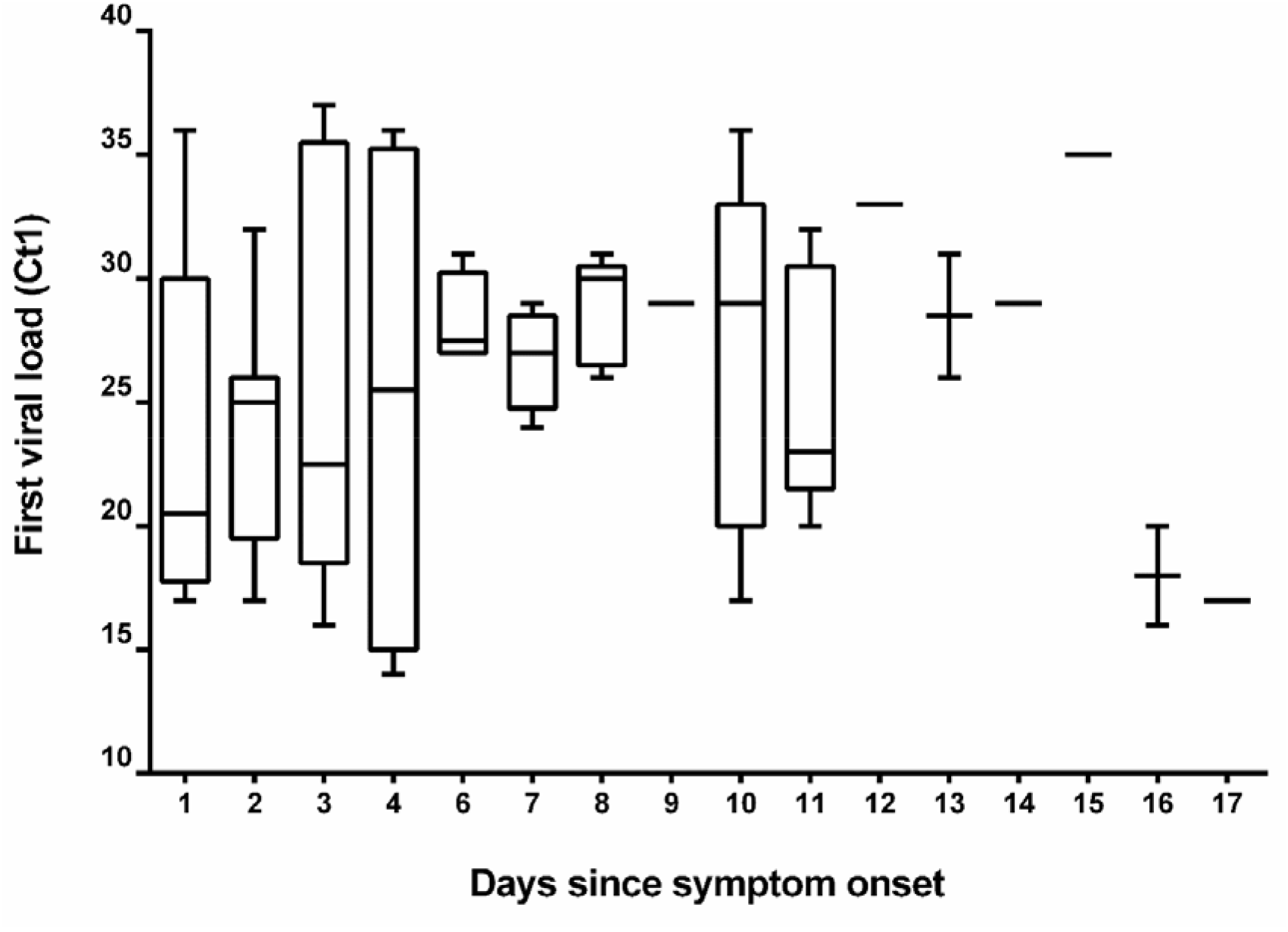
Viral load (Ct values) detected by RT-PCR in the first nasal swabs from patients infected with SARS-CoV-2 (*N*□= □ 66) at hospital admission.

Patients were categorized into two periods according to the timing of sample collection in relation to illness onset. There was no difference in Ct values obtained from the first samples (*p*=0.335), demonstrating the results are homogeneous among the values of the first sample (Table 1).

**Table 1:** Timing of sample collection and RT-PCR (Ct) results.

| Days | N | Average | SD* | *p*-value |
| --- | --- | --- | --- | --- |
| 1 - 7 | 39 | 25.10 | 6.56 | 0.335 |
| 8 or more | 27 | 26.63 | 5.85 |  |

\*SD, Standard deviation

We compared the median viral load obtained for different clinical stages of SARS-CoV-2 infection (Ct1, Ct2, and Ct3 values) between the group of patients who underwent HCQ treatment in comparison with a control group that did not receive the drug. For such comparison, Student’s T-test was used to analyze Ct1 values of first samples at hospital admission, comprising all patients prior to the HCQ administration as an optional drug for treatment. On the other hand, the Mann-Whitney test (for non-parametric data) was used to analyze the Ct2 and Ct3 value differences between patients that have used HCQ in comparison to the control group (no HCQ use). The data are shown in table 2.

**Table 2:**
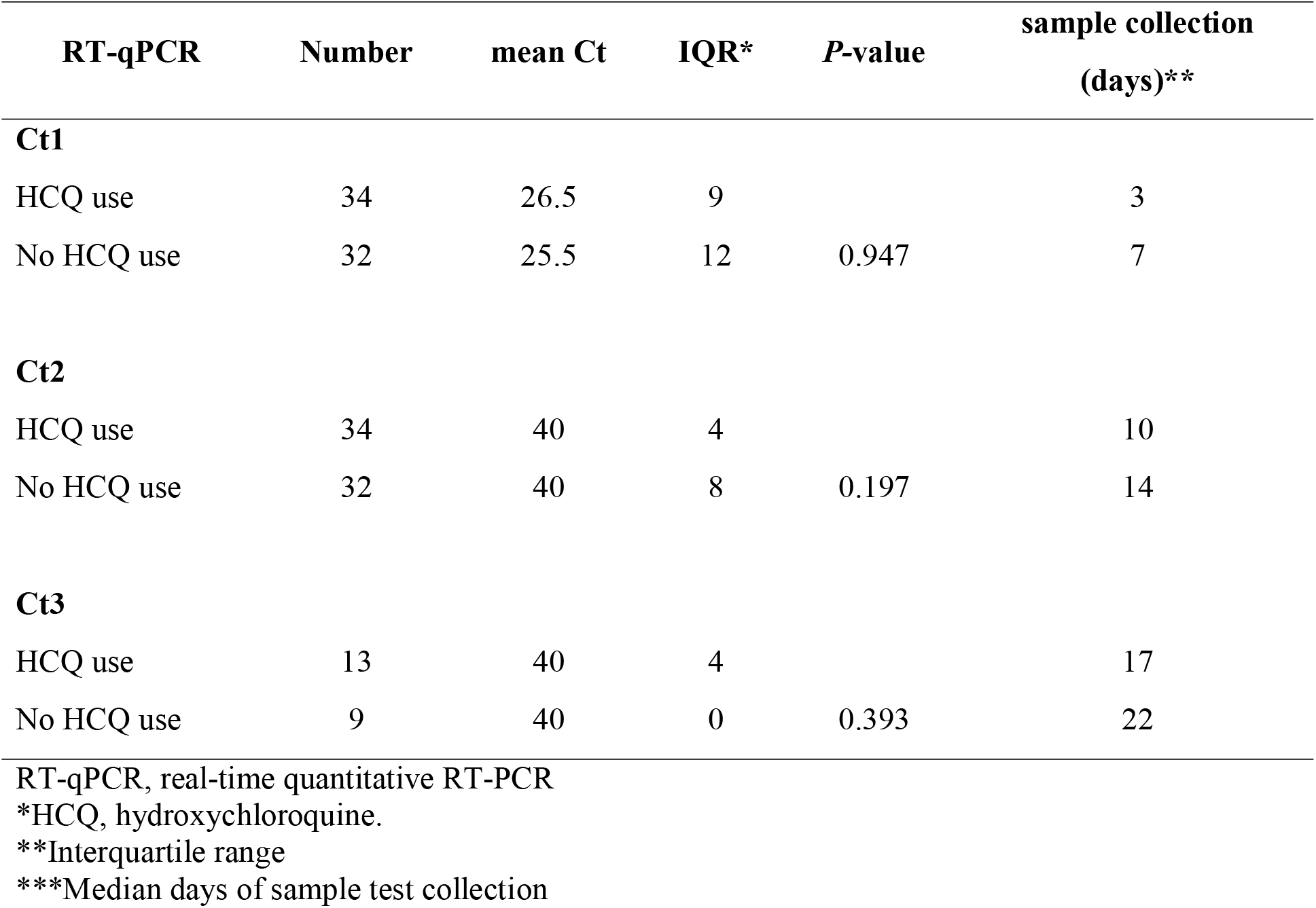
Ct values over time according to HCQ* therapy.

Next, we compared patients with use and no use of HCQ according to clearance of viral load (ΔCt) at intervals between the first and second sample collection (Δt). The two groups were analyzed on the three intervals defined by data of all subjects sampling over the study. No significant viral load clearance was observed among the groups (*p*=0.362 for Δt<7 days, 0.403 for Δt 7-12, and 0.516 for Δt>12 days, two-way ANOVA test). The data are shown in figure 2.

**Figure 2:**
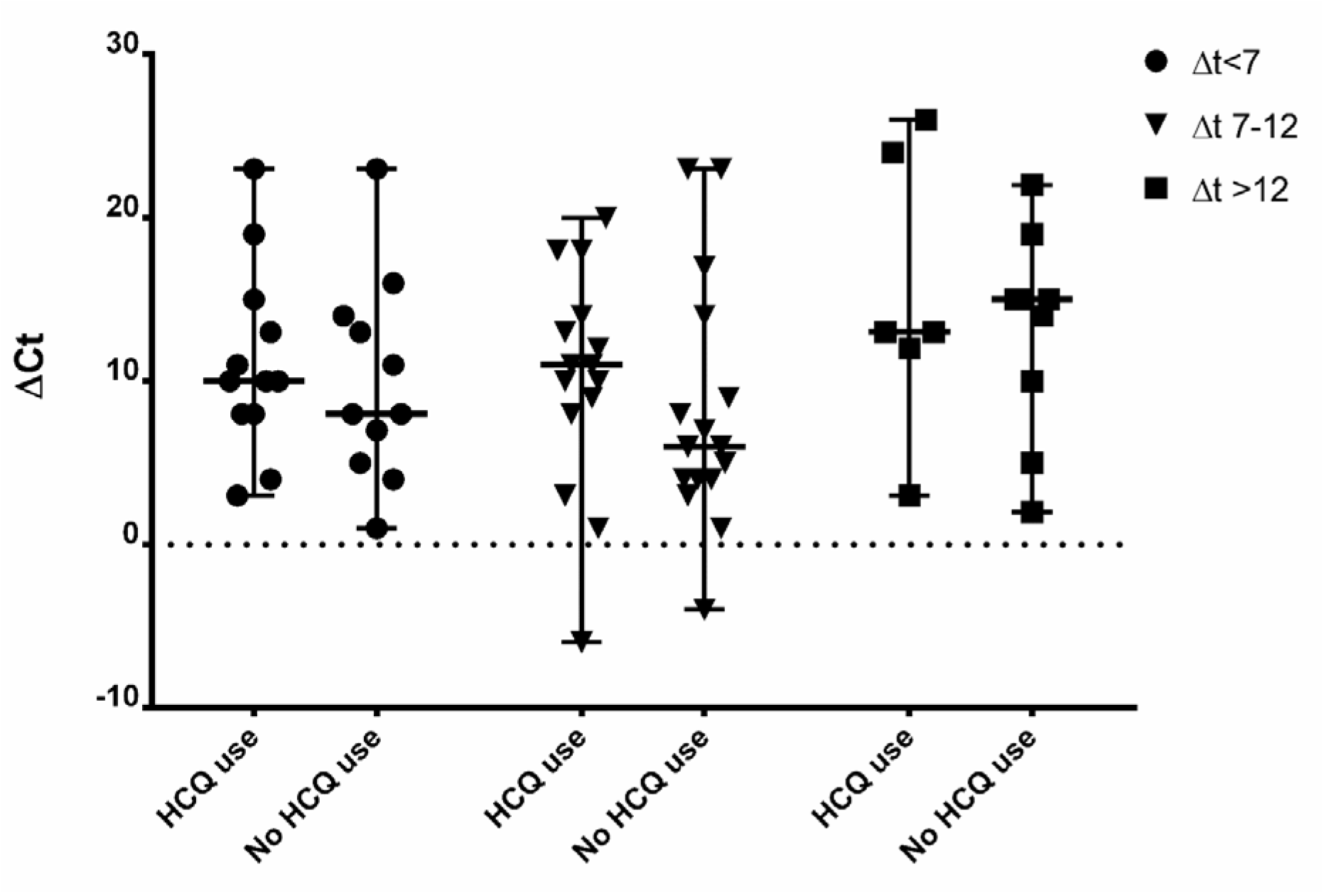
ΔCt among groups (use and no use of HCQ) in different Δt (<7, 7-12, and >12 days). Upper and lower bars indicate min. and max. ΔCt values. Horizontal bars indicate the mean ΔCt.

## DISCUSSION

The hypothesis that HCQ could be effective is still open. Although the study with a greater number of patients did not show differences in outcome [2], the theme remains under discussion even though the fact that their effect on viral replication is not well understood.

We establish a study to assess viral clearance in patients without a severe outcome so that we could compare patients more homogeneously. The viral load is an established method used for the evaluation of therapies and was a biomarker on the study of lopinavir/ritonavir [14]. Viral load clearance also was used in many studies for SARS-CoV-2 showing a gradual decrease in tertiary patients [15, 16].

Zou et al. [16] reported that patients with a severe form of COVID-19 who required hospitalization in an intensive care unit had a high viral load 10 days after the onset of respiratory symptoms and later as well. Lescure et al. [17] reported the viral load dynamics of two patients who later developed respiratory deterioration despite the disappearance of nasopharyngeal viral RNA. They also suggested that the viral load could be used to indicate possible clinical strategies for the treatment of COVID-19.

We could not find any difference in viral load reduction among samples of patients considering the same time score established in other studies that analyzed the clearance of viral load in different clinical presentations. Liu et al. [18] in their study analyzed the clinical outcome of COVID-19 subjects but none patient was under HCQ.

The results of this study must be interpreted in light of methodological limitations. All patients included were hospitalized with moderate severity neither critical nor mild and the difference in viral load may be less striking than a large inclusion could demonstrate. In addition, it is possible that measuring viral load in the nasopharynx may not fully capture the total amount of virus shed by an individual. Patients were selected based on the availability of samples that had tested positive for COVID-19. Therefore, the sampling time of all serial samples for each patient presented some variation. To reduce any bias, we categorized all patients using the ΔCt and Δt analysis to minimize that limitation.

In conclusion, we did not observe any change in viral load *in vivo* with the use of HCQ.

## Data Availability

None

## ACKNOWLEDGMENT

L.K.S.L and J.M.A.C. are a fellow of the Coordenação de Aperfeiçoamento de Pessoal de Nível Superior (CAPES), Brazil. D.D.C. is a fellow of the Conselho Nacional de Desenvolvimento Científico e Tecnológico (CNPq), Brazil.

